# Scrutiny for Child Abuse and Neglect During the COVID-19 Pandemic

**DOI:** 10.1101/2020.10.12.20210997

**Authors:** Maryaline Catillon, Kenneth D. Mandl

**Author notes:** Corresponding author: Kenneth D. Mandl,.

## Abstract

The extant infrastructure for child abuse surveillance, dependent on reporting by schools and healthcare professionals, has been disrupted by the pandemic. Using Google Trends and MediaCloud data, we find a drop in Internet searches and news reports about child abuse and neglect during the pandemic, which may reflect decreased scrutiny.

## INTRODUCTION

The extant infrastructure for surveillance of child abuse and neglect, highly dependent on reporting by schools and healthcare professionals, has been disrupted by the pandemic. Despite concerns that child abuse and neglect might be surging during the COVID-19 pandemic,^1,2,3^ reports of maltreatment have dropped.^4,5^ We sought to quantify the level of public scrutiny for child abuse and neglect, before and during the pandemic, by measuring related Internet searches and news reports as a proxy.

## METHODS

We compare the period of January 1, 2020 through July 31, 2020, to baseline averages from the same period over the five prior years, from 2015 to 2019. US-based Internet search volume was measured using Google Health Trends, with queries for “child abuse” or “child neglect.” The metric is a multiple of the share of U.S. Google searches meeting our query conditions among all Google searches on a given date. News volume was measured using the 225 news sources in MediaCloud’s US National Corpus. Queries of the news sources were for all articles mentioning “child abuse” or “child neglect.” News volume is the ratio of news meeting our query conditions among all US news reported in MediaCloud’s US National Corpus on a given date. Both data sources were normalized so that the average of the five-year baseline in January equals 100.

## RESULTS

During the baseline years (2015-2019), Internet search volume pertaining to child abuse and neglect exhibits strong seasonality. Volumes peak in April and drop off at the end of the school year in June. The pattern observed in 2020 is different. Prior to the social distancing measures implemented for the pandemic, in January and February 2020, search volume was close to the levels observed in the five prior years. However, search volume dropped dramatically between March and May 2020 returning to pre-pandemic levels in June, at the end of the school year (Figure 1).

**Figure 1.**
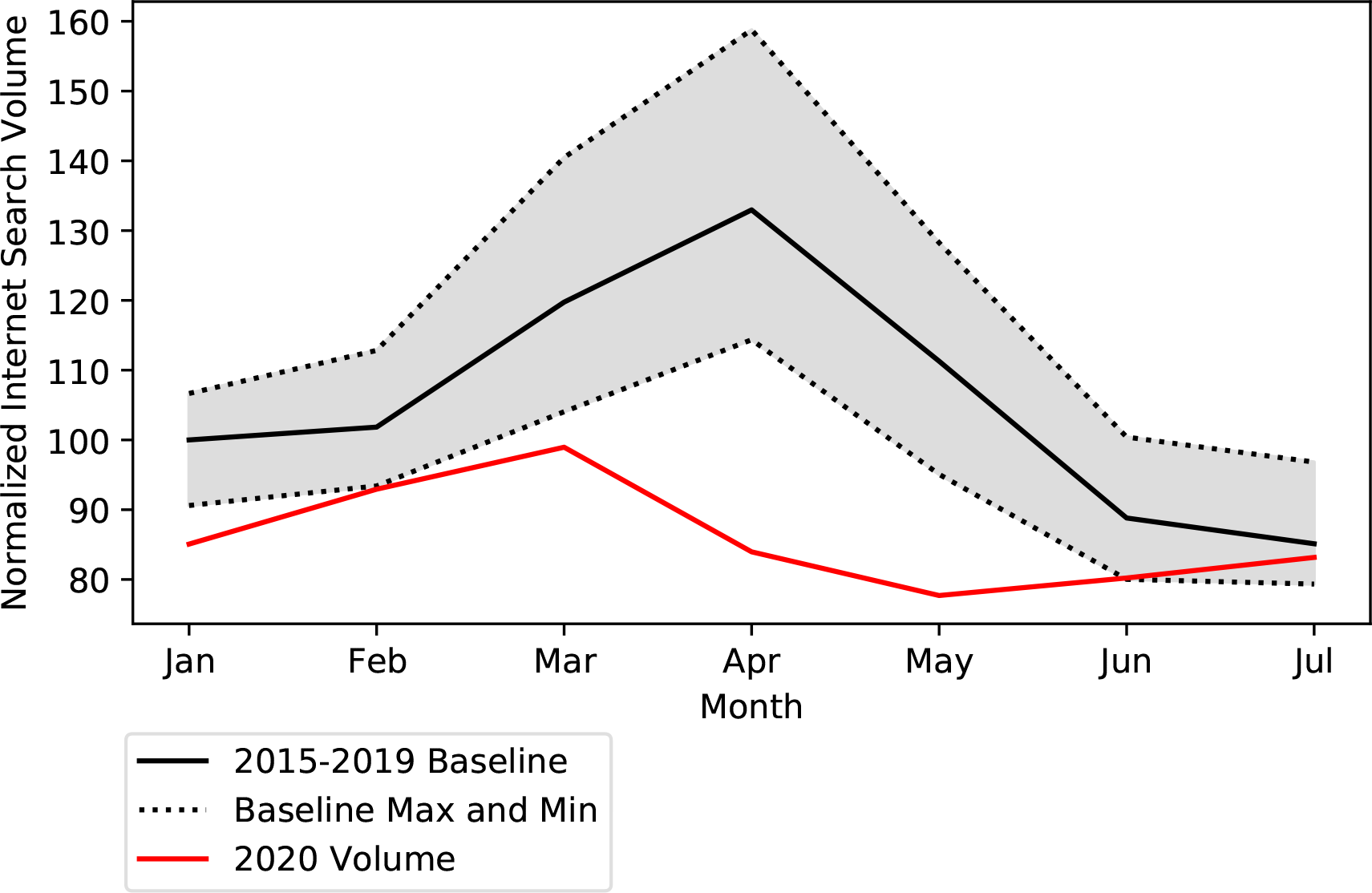
Average Internet search volume for ‘child abuse’ or ‘child neglect’ during the five-year baseline (2015-2019) and Internet Search Volume in 2020. The gray area represents the range of volumes.

In January and February 2020, news volume pertaining to child abuse and neglect was similar to the monthly volumes observed during the prior five-year baseline. However, from March to July, volume was consistently lower than five-year baseline (Figure 2).

**Figure 2.**
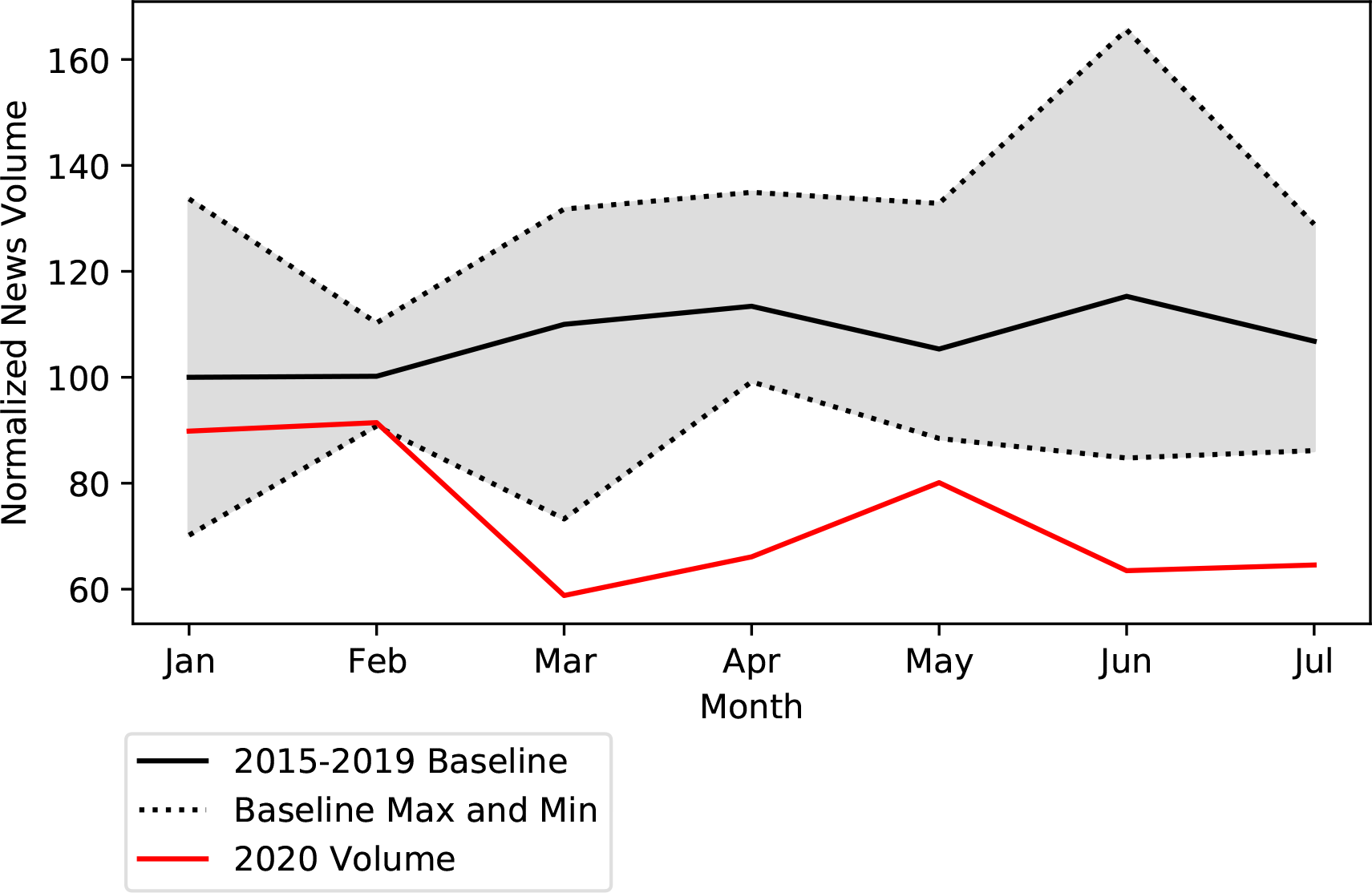
Average news volume for ‘child abuse’ or ‘child neglect’ during the five-year baseline (2015-2019) and news volume in 2020. The gray area represents the range of volumes.

## DISCUSSION

Decreased volumes of Internet searches for child abuse and neglect may reflect inadequate scrutiny of child safety during the pandemic. Schools going online and major disruptions in the healthcare delivery system have reduced contacts between children and the professionals most likely to report child abuse and neglect. Lower news volumes may be a result of fewer high profile cases being identified, but also may contribute to decreased awareness of child abuse in the population and negatively affect detection and reporting. Both an increase in child maltreatment and a decrease in referrals have been previously observed during economic downturns.^6^

This study is observational and relies on a proxy to measure scrutiny for child abuse. Further research is needed to study the relationship between the scrutiny proxy, child abuse reports, and objective measures of maltreatment rates.

The resounding silence in news and search trends should in no way lead to complacency. Quite the opposite; recalling Arthur Conan Doyle’s famous Sherlock Holmes clue to a mystery, the dog that did not bark, we should be redoubling our efforts to find and rescue children at risk, particularly as many schools are beginning online in the 2020-2021 school year.

## Data Availability

The data is available from Google Health Trends and MediaCloud.

